# Greenspace, air pollution, and respiratory health outcomes: A systematic review of cohort studies

**DOI:** 10.1101/2025.10.16.25338155

**Authors:** Mae Vance, Tyler Lane, Tingting Ye

**Affiliations:** School of Public Health and Preventive Medicine, Monash University, Melbourne, VIC 3004, Australia

**Keywords:** Greenspace, air pollution, particulate matter, respiratory health, NDVI

## Abstract

Exposure to greenspace is associated with improved health outcomes, with proposed mechanisms including improved local air quality and health effects that reduce personal susceptibility to air pollution. This systematic review synthesises cohort study evidence on whether greenspace protects against air pollution effects on respiratory-related health outcomes. We conducted a systematic review of cohort studies, searching Medline, Embase, and Scopus databases published up to December 2024. Study quality was evaluated using the Newcastle-Ottawa Scale. Twelve studies met the inclusion criteria, all of which were rated as “good” quality. Cohorts mostly originated from Europe, with outcomes including respiratory-related mortality, lung function, and non-infectious respiratory diseases. In general, exposure to greenspace was protective against air pollution-associated respiratory mortality, with effects more consistently detected through mediation than moderation. Residential greenspace appears to reduce the harm associated with air pollutants on respiratory health outcomes, including related mortality. The stronger association detected through mediation analysis suggests that the mechanism is through improvements in local air quality rather than the direct effects of greenspace on health to enhance individual resilience to the effects of air pollution.

**Highlights:**

- This systematic review investigated whether greenspace protects against respiratory health harms from air pollution
- We identified 12 eligible cohort studies, all rated "good" quality
- Results mixed but generally suggested greenspace is protective
- More consistent findings for mediated pathways, suggesting greenspace reduces pollution in local environment

**Visual Abstract:** 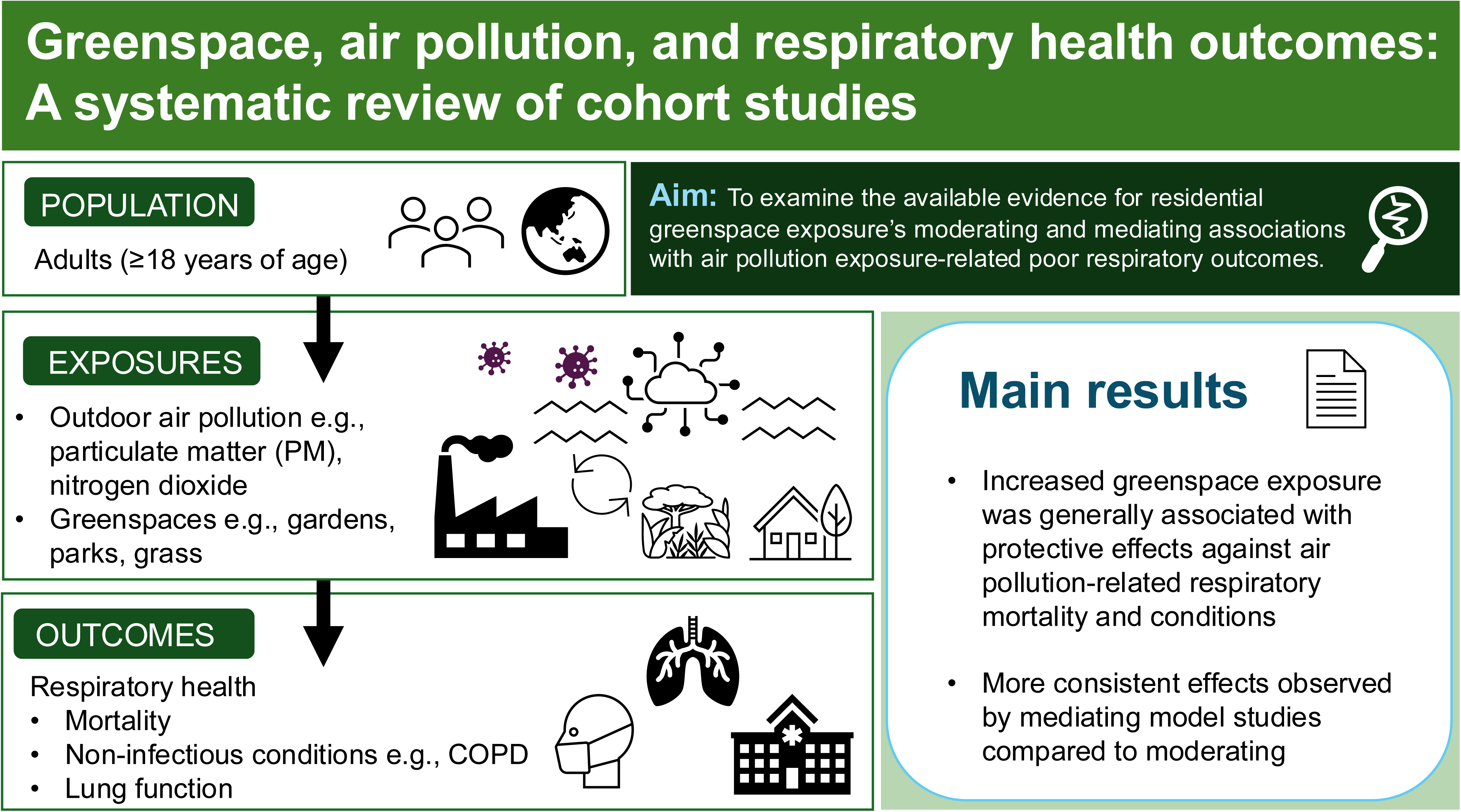

## 1 Introduction

Ambient outdoor air pollution contributes to 7.6% of mortality and 4.2% of disability-adjusted life-years worldwide [1]. Polluting toxins accumulate in lower and upper airways, affecting alveolar tissue and translocating systemically [2]. Unsurprisingly, they are strongly associated with inducing or exacerbating existing conditions, including asthma [3], chronic obstructive pulmonary disease (COPD) [4], and lung cancer [5]. Exposure to air pollution is pervasive globally and unequally distributed [6]. Identifying environmental factors that could mitigate the effects of air pollution could offer insights as to how local communities can be modified to improve community health and are therefore of immense value. Greenspace containing natural and built environments, such as parks and gardens, are one potential mediator or moderator of air pollution [7, 8].

Air pollution originates from both natural and anthropogenic sources such as traffic emissions and pollen spores, and include particulate matter (diameters of <1µm, <2.5µm, and <10µm, or PM_1.0_, PM_2.5_, PM_10_), carbon monoxide (CO), carbon dioxide (CO_2_), methane, and ozone (O_3_) [9, 10]. During wildfires and other extreme weather events, hydrocarbons and oxygenated organic molecules, trace elements, free radicals, and other constituents may also release into the atmosphere [11]. PM_2.5_ is considered one of the most harmful pollutants due to its small size, which enables particles to penetrate the lungs and cause systemic inflammation and decrease immune function [12].

Several non-mutually exclusive mechanisms have been proposed to explain how greenspace might mitigate the health effects of air pollution. Vegetation can alter the chemical composition of pollutants as they deposit on leaf surfaces and reduce ambient temperatures [13]. Tree coverage can reduce windspeed, minimising pollutant dispersion [14]. By blocking UV rays, foliage can lower temperature and the formation of ozone (O_3_) [15]. Through the diffusion of gases and absorption of pollutants [8], plants may remove heavy metals and polycyclic hydrocarbons (PAHs) [16] before they are washed away by rain, fall to the ground, or resuspended into the atmosphere [14]. Additionally, greenspace may be able to directly improve health by minimising psychological stress and encouraging physical activity [17], thus increasing resilience to the harmful effects of air pollution.

There is accumulating research on how greenspace may protect against air pollution’s effects on respiratory health, as indicated by a 2021 systematic review [18]. In this review, we provide more up-to-date review focusing specifically on respiratory health and higher-quality study designs to address the following research question: Does greenspace moderate and/or mediate the impact of air pollutants on respiratory health?

## 2 Methods

This systematic review was registered on PROSPERO, with open access to its study protocol made available on the 5^th^ of December 2024 [19]. Methods and findings are reported according to PRISMA 2020 guidelines [20].

### 2.1 Eligibility Criteria

This review focused on cohort studies where the outcomes of interest were respiratory-related symptoms, non-communicable diseases, and mortality, correlated with degrees of exposure to air pollutants as moderated or mediated by greenspace. Eligibility criteria are presented in Table 1.

**Table 1.** Inclusion and exclusion criteria.

| Category | Inclusion | Exclusion |
| --- | --- | --- |
| **Conditions** | Respiratory function, symptoms, non-infectious conditions such as asthma and COPD | Respiratory-related cancers and communicable diseases |
| **Exposures** | Outdoor air pollutants, including PM<sub>2.5</sub>, and any measured greenspace | Indoor air pollution |
| **Populations** | Adults (18 ≥ years of age), living in metropolitan or rural areas | Children (<18 years of age) |
| **Study designs** | Cohort studies | Cross-sectional, case-control, case-cross-over, randomised controlled trials, and ecological studies |

### 2.2 Search strategy and screening

We systematically searched the following journal databases for relevant articles: Ovid-Medline, Ovid-Embase, and Scopus. The search strategy (Table S1 in Supplementary Materials) was developed with reference to previous systematic reviews on the topics of greenspace, respiratory health [21], and wildfire smoke [22]. Search results were not restricted by date of publication.

### 2.3 Data extraction and quality assessment

Search results were transferred into Covidence [23] and screened. Two reviewers (MV, TY) independently screened titles, abstracts, and full texts, with disagreements resolved through discussion. Two reviewers (MV, TL) assessed quality using the Newcastle-Ottawa Scale (NOS) [24]. We selected socioeconomic status or any other proxy (e.g., education, employment, ethnicity) as the most important confounder to control for. Instead of satisfying the criterion that the outcome of interest was demonstrated not to be present at the start of the study, we awarded a star if the study tested for the outcome at baseline.

## 3 Results

### 3.1 Search results and study characteristics

The screening process is outlined using the PRISMA flow chart, as shown in Figure 1. After removing duplicates, 770 articles’ titles and abstracts were screened, of which 733 (95%) did not meet the inclusion/exclusion criteria. Full-text screen excluded a further 25 studies due to ineligible design or non-English publication. This resulted in 12 included studies.

**Figure 1.**
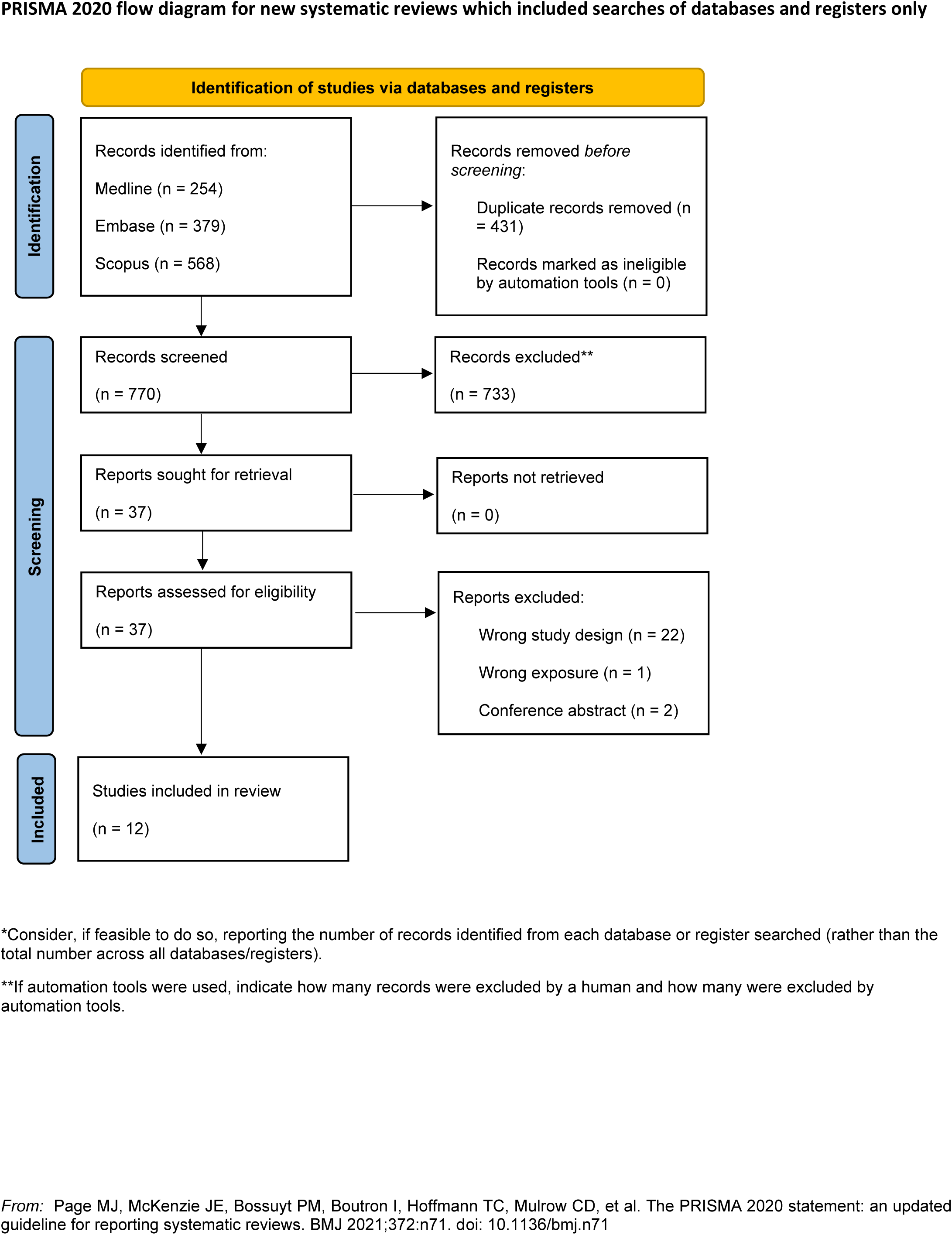
PRISMA 2020 flow diagram, adapted from Page et al. [20]

Most study cohorts came from European settings (*N* = 6, 50%), followed by China (*N* = 4, 33%), and one each (8%) in the USA and Korea. A variety of respiratory-related health outcomes were measured across these studies, most often mortality (*N* = 7, 58%), including COPD-related mortality (*N* = 2). All studies had retrospective cohort designs, and most utilised data linkage to measure associations between respiratory-related outcomes, air pollutants, and greenspace exposures. The same cohort was used in multiple studies, including two from UK Biobank [25, 26] and four from the Chinese Community-based Collaborative Innovation Hepatitis B Virus (CCI-HBV) cohort [27–30].

Greenspace measurement varied across and within cohorts. Most studies assessed greenspace exposure using the *Normalised Difference Vegetation Index* (NDVI) from satellite imagery, testing multiple buffer radii (i.e., width of the circle around the residential geocoordinates). One study additionally incorporated land-use data to denote agricultural and natural greenspace such as forests in proximity to residences [31].

An overview of the included studies’ characteristics, grouped by outcome, can be found in Table 2.

**Table 2.**
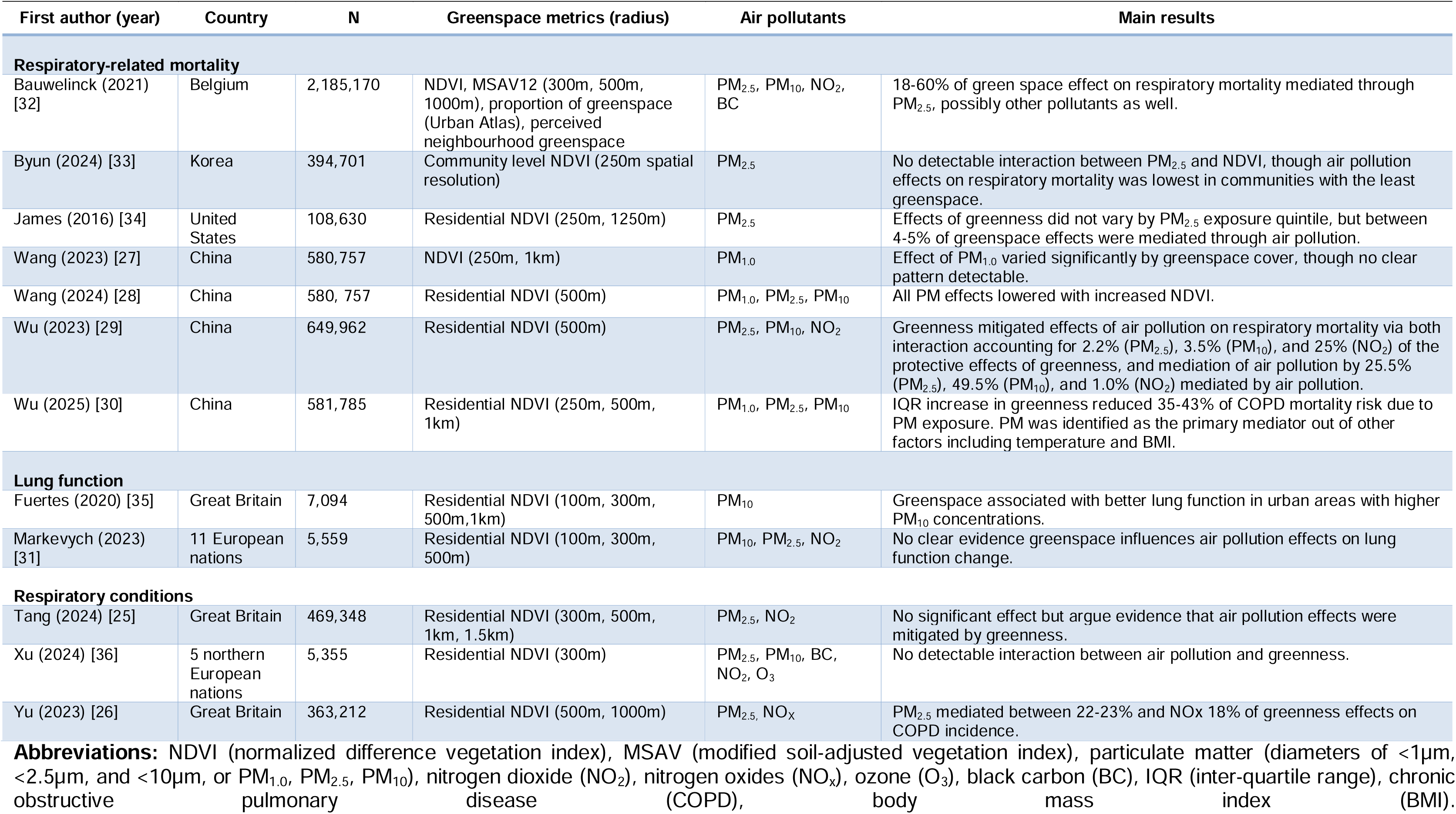
Summary of included studies.

### 3.2 Study quality

Quality assessment scores are presented in Table 3. The quality of studies is indicated by the number of stars or points within each domain (Selection, Comparability, Outcome). All were classified as “good quality”, the highest rating, based on conversion of NOS scores to AHRQ standards. All studies received a star for “selection of the non-exposed cohort” (each used continuous exposure metric, forgoing the need to recruit “unexposed” individuals) and “ascertainment of exposure” (each used satellite-generated images of greenspace and modelled air pollution data linked to residential address) in the Selection domain. For the Comparability domain, each study received the maximum two stars because they adjusted for at least one socioeconomic status indicator and at least one other control variable.

**Table 3.** Data quality and risk of bias assessment results for included cohort studies (*N* = 12) based on Newcastle-Ottawa Scale (NOS) for cohort studies [24].

| Study | Domain and score |  |  |
| --- | --- | --- | --- |
|  | Selection | Comparability | Outcome |
| Bauwelinck et al. (2021) [32] | ☆☆☆☆ | ☆☆ | ☆☆☆ |
| Byun et al. (2024) [33] | ☆☆☆— | ☆☆ | ☆☆☆ |
| Fuertes et al. (2020) [35] | ☆☆☆☆ | ☆☆ | ☆☆— |
| James et al. (2016) [34] | ☆☆☆— | ☆☆ | ☆☆☆ |
| Markevych et al. (2023) [31] | ☆☆☆☆ | ☆☆ | ☆☆— |
| Tang et al. (2024) [25] | ☆☆☆— | ☆☆ | ☆☆— |
| Wang et al. (2024) [28] | ☆☆☆☆ | ☆☆ | ☆☆☆ |
| Wang et al. (2023) [27] | ☆☆☆☆ | ☆☆ | ☆☆☆ |
| Wu et al. (2023) [29] | ☆☆☆☆ | ☆☆ | ☆☆☆ |
| Wu et al. (2025) [30] | ☆☆☆☆ | ☆☆ | ☆☆☆ |
| Xu et al. (2024) [36] | ☆☆☆☆ | ☆☆ | ☆☆— |
| Yu et al. (2023) [26] | ☆☆☆— | ☆☆ | ☆☆☆ |

Most of the studies were sizeable, including several hundred thousand participants, while one included over 2 million [32]. However, three studies had under 10,000 participants each [31, 35, 36].

There were nevertheless some limitations uncaptured by the NOS. The lingering threat of immortal time bias due to mortality effects of both greenspace and air pollution before study follow-up can begin was only overcome by Fuertes et al. [35], who recruited participants *in utero* (via their pregnant mothers). Studies relying on residential address provided at recruitment, including those using UK Biobank data and the CCI-HBV data did not account for variations in greenspace and air pollution due to subsequent change of address. All others updated these. One study included several outcomes for COPD (diagnosis, hospitalisation, mortality) but only evaluated greenspace’s protective effects against air pollution for mortality [30].

### 3.3 Evidence synthesis

All studies included PM, usually PM_2.5_, as an air pollution indicator; other air pollution indicators included nitrogen oxides (NO_X_), nitrogen dioxide (NO_2_), black carbon (BC), and Ozone (O_3_).

#### 3.3.1 Respiratory-related mortality

Of the 7 studies investigating respiratory-related mortality, 4 found that greenspace protected against air pollution [28–30, 32], 1 found null or potentially harmful effects [33], 1 found mixed null and potential protective effects [34], and 1 claimed protective effects while plotted results indicated no clear pattern [27].

Two studies, both using the same Chinese cohort, focused on COPD mortality. One reported that “participants with lower NDVI [greenness] exposure were more susceptible to the influence of PM_1_ than those with higher NDVI exposure” [27]. However, the plotted results suggested similar effects between the top and bottom greenness quartiles, indicating the effect was not consistent. The other study [30] used a multiple mediation analysis to investigate both interaction and mediating effects between greenspace and PM of varying sizes, detecting both.

Aside from Wu et al. [30], effects were infrequently detected in interaction or stratified analyses (assuming a multiplicative effect between air pollution and greenspace) but were frequently detected in mediation analysis (assuming greenspace’s mechanism is reducing air pollution). For instance, the American study found no interaction between ambient PM_2.5_ and greenspace, but 4-5% of greenspace’s protective effects against respiratory mortality were mediated through air pollution [34].

A Belgian study [32] observed both greater surrounding greenspace and greater perceived greenspace were associated with lower respiratory mortality due to ambient PM_2.5_ and potentially other air pollutants as well. The authors suggested that perceived neighbourhood greenspace may have accurately captured aspects of greenspace relating to its moderating effects, including walkability and biodiversity. Results varied across the different types of greenspace exposure and pollutant types; more so for residential surrounding greenness (NDVI) and residential surrounding greenspace (Urban Atlas), than for perceived neighbourhood greenspace.

One Chinese study [29] was the only one to find both moderating and mediating effects of greenness on air pollution (PM_2.5_, PM_10_, and NO_2_). Another Chinese study [28] found the effects of PM air pollution were greatest in the lowest tertile of greenspace areas; areas with more NDVI were theorised to encourage greater exercise, thereby directly improving health outcomes. In contrast, a South Korean study found no association between PM_2.5_ and respiratory mortality among those living in the lowest tertile of community-level greenspace, but it was associated with *increased* mortality in the middle and highest tertiles [33]; the authors suggested several aspects of greenspace that could exacerbate air pollution effects: seasonal release of allergenic pollens, use of pesticides, dense vegetation increasing risk of vector-borne disease, and the formation of secondary organic aerosols and ground-level ozone.

#### 3.3.2 Lung function

Two studies investigated lung function using spirometry [31, 35]. In a British cohort of younger individuals, Fuertes et al. [35] reported that the association between urban greenspace and better lung function (FEV_1_ [forced expiratory volume in one second] or capacity and FEF [forced expiratory flow] or small/narrowing airways) was higher in areas with greater ambient PM_10_ (the only pollutant tested). In contrast, a study of young to middle-aged adults across 11 European nations [31] found no consistent evidence between types of air pollution (NO_2_, PM_2.5_, PM_10_) and modification of greenspace effects on lung function (FEV_1_ and FVC or indicator of airway obstruction or disease).

#### 3.3.3 Respiratory conditions

Three studies investigated respiratory conditions, including two of COPD [26, 36] and one of idiopathic pulmonary fibrosis [25]. The studies of COPD were mixed. Xu et al. [36] used data from participants in 5 European countries and tested multiple air pollutants (PM_2.5_, PM_10_, BC, NO_2_, O_3_), none of which elicited an interaction with greenspace. In contract, Yu et al. [26] used data from UK Biobank and found both PM_2.5_ and NO_X_ (the only air pollutants tested) both mediated greenness effects on COPD, regardless of buffer size. Differences in findings may be partly attributable to study power: Yu et al. [26] analysed over 360,000 participants with more than 8,000 COPD cases, compared with just over 5,000 participants and 328 cases in Xu et al. [36].

The study of idiopathic pulmonary fibrosis (IPF) used UK Biobank data, reported that PM_2.5_ and NO_2_ were the main mediators of greenspaces effects on risk of incident IPF [25]. Mediation effects were stronger at smaller buffer sizes (< 1000m). However, while the effects were all in the same direction across both pollutants and greenspace buffers sizes, confidence intervals were wide and none of the associations achieved statistical significance.

### 3.4 Mechanisms

The included studies adopted a statistical approach that assumed a relationship between greenspace and air pollution that was mediated, moderated, or both. Each entails different mechanisms, though they are not necessarily mutually exclusive. Mediation analysis assumes greenspace affects localised levels of air pollution by removing pollutants and reducing dispersion from wind turbulence [26, 29]. Moderation analysis assumes that greenspace does not necessarily reduce pollution levels, but that it changes the strength of the associations between air pollution exposure and health outcomes. The moderating effect is the improved general health of individuals living around more greenspace, e.g., reduced individual stress and increased exercise habits [21].

Across the included studies, mediation analysis was most frequently used, reported in seven studies [25, 26, 29, 30, 32, 34, 35]. Mediation was often tested using structural equation modelling. All but one of the mediation studies [35] detected an effect; Tang et al. [25] also reported a mediating effect of air pollution, though this was not statistically significant.

Moderation analyses, conducted with both interaction terms and stratifying analysis into quantiles of either air pollution or greenspace, were used in five studies [27, 28, 31, 33, 36], and were less consistent. One moderation study even found possible evidence of a harmful effect of greenspace [33], which was noted as being potentially related to unmeasured confounders, including allergenic pollen blooms. One study [29] tested both mechanisms, using a causal mediation estimation and a four-way decomposition method that evaluated both mediation and moderation effects simultaneously, and found evidence for both. Notably, the moderating effects were considerably larger for the gas air pollutant, NO_2_, whereas mediating effects were considerably larger for particulate matter. The only other study to evaluate both mediation and moderation effects [34] did so in separate models, only finding evidence for the former.

## 4 Discussion

Although the findings were not entirely consistent, 8 of 12 studies suggested that greenspace offered some protection against negative respiratory health outcomes in the context of air pollution. This was most evident for respiratory-related mortality, where mediation effects were more consistently observed than moderation effects. While the number of eligible studies was limited, they collectively included more than 3.8 million participants, with individual sample sizes ranging from 5,355 to over 2.18 million. The distribution of outcomes was uneven, with seven studies examining respiratory-related mortality, two investigating lung function, and three assessing respiratory conditions. While we identified some methodological issues, all studies were rated as “good” quality.

The results of this review align with a 2021 systematic review [18], which found that residing in areas with more greenspace was associated with a smaller association between PM_2.5_ and respiratory-related mortality and hospitalisation. Our review builds on the previous review by incorporating more recent evidence and comparing findings from moderating and mediating analyses.

Results from the mediation analyses were more consistent than moderation, providing greater support for this mechanism, whereby greenspace protects against air pollution effects by reducing air pollution. However, this does not necessarily rule out a moderation effect, whereby greenspace protects against air pollution effects by improving health, as these mechanisms need not necessarily be mutually exclusive, as indicated by Wu et al. [29]. Notably, the associations across studies examining greenspace’s meditating effects on respiratory-related mortality did vary considerably across types of pollutants, and radii of greenspaces. In another mortality study, significant protective associations were only found for greenspace’s effects on some pollutants, such as PM_10_ [32]. Despite the variability of findings for greenspace and respiratory-related mortality, the overall direction of associations indicated beneficial effects of greater greenspace exposure.

The somewhat mixture of results across studies may have been partially due to variations in greenspace exposure measurement and NDVI buffer sizes, which ranged from 100 to 1500m. There is some evidence that a 300m NDVI radius may best capture urban greenspace accessible within walking distance [37]. This may support the theory of greenspace’s moderation, or lifestyle, effect on health outcomes. However, as there is not one accepted radius buffer size to capture greenspace’s possible mediation effects (nor may one be feasible for all analytical purposes), most included studies applied various NDVI buffer radii in their analyses. Some studies observed slightly stronger mediation effects in smaller NDVI buffers (e.g., 200m) compared with larger ones (e.g., 1000m) [25, 26, 34]. This may have been due to smaller buffers being able to capture fine-scale variation in greenspaces for participants living near each other. Other potential confounders contributing to heterogeneity across study findings include variation in quality of greenness, air pollution measurements being less precise (i.e., more localised) than greenspace measurements, and the pathology of some conditions. As noted in Xu et al. [36], regarding COPD - factors including temporal onset and latency periods may affect associations with environmental exposures.

Additional consideration relates to confounder adjustment. While all included studies were rated as good quality, the set of confounders controlled for varied across cohorts. Socioeconomic status (SES) or related proxies (e.g., education, employment, ethnicity) were identified as the most important confounders in the associations between air pollution, greenspace, and respiratory health. Not all studies adjusted for SES in the same way. Bauwelinck et al. [32], for instance, applied both individual and area-level SES indicators, including education level, employment status, and area mean net taxable income. Whereas Byun et al. [33] controlled for SES confounding using income and community-level deprivation index tertile variables. Inconsistent or insufficient confounding control across studies may therefore have contributed to the heterogeneity in findings.

Variation in greenspace quality may have affected the strength of associations additionally. As noted by Byun et al. [33], allergenic plants producing pollen may contribute to whether greenspace has a protective or harmful effect.

### 4.1 Strengths and limitations

A strength of this review is the high quality of included studies. Notably, there was consistency across the measurements of greenspace, with each study using residential NDVI radii. Included studies came from regions with both higher (China) and lower (Europe, United States) levels of air pollution.

One of the limitations of this review is the gap in available evidence for the moderation of source-specific types of air pollution, such from as wild or bushfires and specific types of greenspaces on respiratory outcomes. The variation in types of air pollutant exposure, such as by ambient air pollution and extreme fire smoke exposure, should be considered by future researchers [38]. None of the studies found by this review measured fire-sourced air pollutant exposure and greenspace. It is important to consider that levels of pollutant toxicity in similar concentrations may vary significantly from ambient sources to fire smoke [39].

Further, across the included cohorts, the settings, populations, and measurements of individual-level confounding variables may limit the transferability of findings [40]. Additionally, over the study period, covariate information may have changed or been inaccurately recorded, such as BMI and physical activity levels measured at baseline [30]. Community-based cohorts may have had less variation in exposures potentially leading to underestimations of effect as well, or a bias towards the null [30].

Differences in greenspace types contributes significantly to outcomes. For instance, the effects of ‘forest’ greenspace exposure may be protective, whereas greenspaces ‘parks’ and ‘grass’ may increase allergen risk, leading to greater adverse health outcomes [41]. For this reason, it may be assumed that the variations of plant and vegetation types across settings can potentially affect the strength of associations [42]. This may be unaccounted for by some studies, as the use of standardised greenspace measurements, such as NDVI, do not differentiate plant type and height [42]. Further, issues related to greenspace measurement include individual exposure calculation. There is the potential for misclassification for greenspace and air pollution exposure calculated by residential address as it does not account for time spent in other areas such as workplaces and commuting in traffic. Additionally, there was a risk of self-selection bias for residential addresses, whereby people may move to greener areas if they are in poorer health [31]. Unfortunately, the results of this review were too heterogeneous to harmonise and combine for statistical assessments such as Egger’s regression or a funnel plot to formally test for publication bias, which remains a possibility.

## 5 Conclusion

While the results were not entirely consistent across studies, they suggested that greenspace reduces harms of air pollution on respiratory health. While by no means conclusive, the findings were most consistent for the mechanism that greenspace reduces air pollution in the local environment; however, the other mechanism – that greenspace improves health directly, providing some resilience to air pollution’s health effects – may also play a role. Most studies relied on a single measure of greenspace, NDVI; there remains a gap as to the protective effects of different types and measures of greenspace.

## Data Availability

All data produced in the present study and not contained in the manuscript are available upon request to the authors.

## CRediT authorship contribution statement

### Mae Vance

Conceptualisation, Methodology, Writing – original draft, Writing – review & editing, Visualisation. **Tyler Lane:** Conceptualisation, Methodology, Supervision, Writing – review & editing. **Tingting Ye:** Conceptualisation, Methodology, Supervision, Writing – review & editing.

