## Supplementary material for "Greenspace, air pollution, and respiratory health outcomes: A systematic review of cohort studies": Table S1 in Supplementary Materials

### Table S1

#### Greenspace MeSH Terms [21]

(“green space” or greenspace* or greenness or greenery).ti,ab.

(wilderness or “wild land” or “natural land” or “natural environment” or “wild space”).ti,ab.

(“municipal land” or “community land” or “public land” or “open land” or “municipal space” or “natural space” or “open space”).ti,ab.

(park or “municipal park” or “botanic park” or “park access” or “urban park” or “city park” or “park availability” or “public garden” or “natural neighbourhood” or “natural facilities”).ti,ab.

(“vegetation natural” or “belt green” or “wild area” or “trail green” or “natural area*” or “green area*”).ti,ab.

(“built environment” or “urban design” or “recreation resource”).ti,ab.

(grass or forest or shinrin-yoku or “forest bathing”).ti,ab.

(NDVI or “Normali*ed Difference Vegetation Index” or EVI or “Enhanced Vegetation Index” or SAVI or “Soil* Adjusted Vegetation Index”).ti,ab.

#### Respiratory health MeSH Terms [22]

(autoimmun* or pneumoconiosis or respiratory or asthma or pulmonary or COPD or “Chronic Obstructive Pulmonary Disease*”).ti,ab.

#### Air pollution MeSH Terms [22]

Wildfires/

(coal*mine fire* or bushfire* or bush fire* or wildfire* or wild* fire* or forest fire* or wildland fire* or wild land fire* or woodland fire* or wood land fire* or brushfire* or brush fire* or rural fire* or grassfire* or grass fire or vegetation fire* or landscape fire* or particulate matter* or PM25 or PM10 or airborne particle* or air pollution* or air pollutant* or air quality). ti,ab.
